# Platelet expression and reactivity after BNT162b2 vaccine administration

**DOI:** 10.1101/2021.05.18.21257324

**Authors:** Melissa Klug, Olga Lazareva, Kilian Kirmes, Marc Rosenbaum, Marina Lukas, Simon Weidlich, Christoph D. Spinner, Moritz von Scheidt, Rosanna Gosetti, Jan Baumbach, Jürgen Ruland, Gianluigi Condorelli, Karl-Ludwig Laugwitz, Markus List, Isabell Bernlochner, Dario Bongiovanni

## Abstract

SARS-CoV-2 infection induces a coagulopathy characterized by platelet activation and a hypercoagulable state with an increased incidence of cardiovascular events. The viral spike protein S has been reported to enhance thrombosis formation, stimulate platelets to release pro-coagulant factors and promote the formation of platelet-leukocyte aggregates even in absence of the virus. Although SARS-CoV-2 vaccines induce spike protein overexpression to trigger SARS-CoV-2-specific immune protection, thrombocyte activity has not been investigated in this context. Here, we provide the first phenotypic platelet characterization of healthy human subjects undergoing BNT162b2 vaccination.

Using mass cytometry, we analyzed the expression of constitutive transmembrane receptors, adhesion proteins and platelet activation markers in 12 healthy donors before and at five different timepoints within four weeks after the first BNT162b2 administration. We measured platelet reactivity by stimulating thrombocyte activation with thrombin receptor-activating peptide (TRAP). Activation marker expression (P-Selectin, LAMP-3, LAMP-1, CD40L and PAC-1) did not change after vaccination. All investigated constitutive transmembrane proteins showed similar expressions over time. Platelet reactivity was not altered after BNT162b2 administration. Activation marker expression was significantly lower compared to an independent cohort of mild symptomatic COVID-19 patients analyzed with the same platform.

This study reveals that BNT162b2 administration does not alter platelet protein expression and reactivity.

## Introduction

SARS-CoV-2 infection and coronavirus disease 2019 (COVID-19) can lead to a hypercoagulable state with increased incidence of adverse cardiovascular events^1–4^. Over the last months, several studies highlighted thrombocyte activation as a key feature of COVID-19 induced coagulopathy^5–7^. We recently described a dysregulation of platelet expression and reactivity even during mild COVID-19 disease characterized by an upregulation of activation markers as well as of the transmembrane proteins GPIIb/IIIa, GPIX and GPIa, which regulate platelet activation^8^. However, the pathophysiological mechanism responsible for COVID-19 induced coagulopathy and of the increased incidence of cardiovascular events remains unclear.

A recent study reported that the SARS-CoV-2 spike protein S enhances thrombosis formation, stimulates human platelets to release pro-coagulant factors and promotes platelet-leukocyte aggregate formation even in absence of the virus^9^. This direct effect on platelets could explain the sustained platelet activation during COVID-19 and raises safety concerns regarding the newly developed vaccines. In fact, although SARS-CoV-2 vaccines induce spike protein overexpression to trigger SARS-CoV-2-specific immune protection, thrombocyte activity has not been investigated in this context. Nevertheless, all vaccines available passed phase III trials^10–12^ but the recently reported thrombotic events following vaccine administration, including BNT162b2, raised concerns worldwide^13,14^. Moreover, an immune thrombotic thrombocytopenia mediated by platelet-activating antibodies against PF4, which mimics autoimmune heparin-induced thrombocytopenia, has been reported after ChAdOx1 vaccine administration^15^.

In this study, we performed the first phenotypic thrombocyte characterization in healthy donors undergoing BNT162b2 vaccine administration using mass cytometry by time of flight (CyTOF).

## Material and methods

### Study design and participants

We recruited 12 healthy donors undergoing vaccination with BNT162b2 without pre-existing conditions or therapy. In all donors, the second dose of BNT162b2 was delivered 21 days after the first one. Blood samples were collected 1 day before vaccination (timepoint 0: TP0) and at 5 different timepoints after the first dose (Figure 1A, supplemental figure 1): day 1-2 (timepoint 1: TP1); day 3-4 (timepoint 2: TP2); day 14 (timepoint 3: TP3); day 22 (one day after the second dose, timepoint 4: TP4) and day 28 (+7 days after the second dose, timepoint 5: TP5). Peripheral venous blood was collected from each donor and immediately processed to platelet-rich plasma and stained for CyTOF analysis as described previously^8,16^. All participants gave written informed consent. The study was approved by the local ethics committee (approval number 4/21 S-KH) and complies to the Declaration of Helsinki.

**Figure 1:**
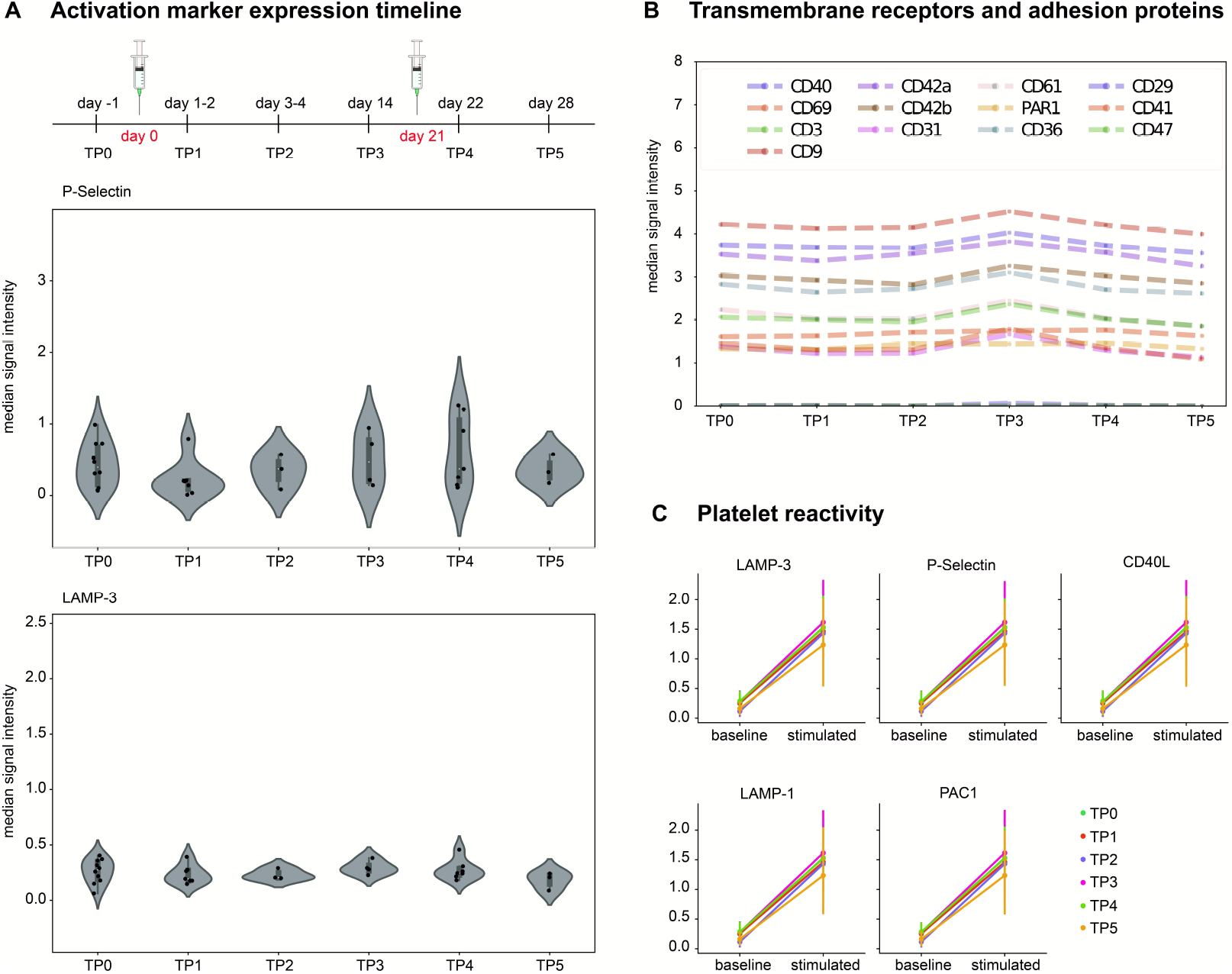
Receptor expression and platelet reactivity after BNT162b2 vaccine administration. A) Expression of activation markers P-Selectin and LAMP-3 over time in non-stimulated samples. Each dot represents the median signal intensity of one donor. Changes in expression were found insignificant between timepoints (Kruskal-Wallis test, supplemental table 3). TP0 n=10, TP1 n=8, TP 2=3, TP 3 n=4, TP4 n=7, TP5 n=3. B) Median expression of constitutive transmembrane proteins over time in baseline samples. C) Activation marker expression (P-Selectin, LAMP-3, LAMP-1, CD40L, PAC1) of baseline samples compared to stimulated samples before and after vaccination.

### Sample collection and preparation

Briefly, two samples were prepared from each blood collection: one baseline sample (non-stimulated platelets) and one sample stimulated with 10 μM thrombin receptor-activating peptide (TRAP). Platelets were stained with a custom-made CyTOF panel of 18 antibodies for 30 minutes (containing anti-CD3-170Er, anti-CD9-171Yb, anti-CD29-156Yb, anti-CD31-145Nd, anti-CD36-152Nd, anti-CD40-142Nd, anti-CD41-89Y, anti-CD42a-141Pr, anti-CD42b-144Nd, anti-CD47-209Bi, anti-CD61-146Nd, anti-CD62P-161Dy, anti-CD63-150Nd, anti-CD69-162Dy, anti-CD107a-151Eu, anti-CD154-168Er, anti-PAC1-155Gd and anti-Par1-147Sm antibodies; see major resources table for antibody information). Samples were then prepared according to the Maxpar cell surface staining protocol as described previously^8^.

### CyTOF acquisition and processing

For acquisition, cells were diluted to a final concentration of 10^3^ platelets/µl in 10% EQ calibration beads in cell acquisition solution (CAS). All samples were prepared and measured by the same scientist. Throughout the study, all samples of one donor were measured at the same day to reduce batch effect. In total, 299,408 ± 1,364 events per sample were acquired at a rate of 200-350 events per second. After acquisition at the Helios CyTOF system (Fluidigm), samples were normalized, processed and pre-gated using the Cytobank™ software (www.cytobank.org, Beckman-Coulter, Brea, CA, USA)^17^.

### Computational Analysis and clustering analysis with FlowSOM algorithm

We performed computational analysis with Cytobank and statistical tests were performed on the transformed data using the scipy python package (version 1.6.3)^18^. For statistical analysis, we used Kruskal-Wallis test for multi-group comparison and Mann–Whitney U test for one-sided pair-wise comparison of different time points. P-values are reported uncorrected for multiple hypothesis testing on the account that no significant p-values were obtained.

FlowSOM analysis was performed using the implementation in Cytobank. Gated, compensated, scaled and transformed files were used as input. 11 markers were used for clustering excluding activation markers, the extremely lowly expressed marker CD40 and the control marker CD3. Cells were assigned to a 10□×□10 self-organizing map before meta-clustering. FlowSOM analysis was run separately for each timepoint and condition using the same minimum spanning tree and seed with 10 000 events per sample. For each cluster, a P value was computed with the one-sided Mann–Whitney U test (see supplemental table 4).

### Comparison with independent cohorts

Protein expression level of the activation markers P-Selectin and LAMP-3 were compared with those of two independent cohorts analyzed previously with the same platform^8^: a group of eight patients with no pre-existing conditions requiring hospitalization due to symptomatic COVID-19 and a control group of eleven healthy donors tested negative for SARS-CoV-2 infection.

### Data and Code Availability

All mass cytometry data and FlowSOM results have been made available at flowrepository.org^19^ and can be accessed at repository ID FR-FCM-Z3XS. The scripts used in this analysis have been deposited at github.com and can be accessed at https://github.com/biomedbigdata/Platelet-activation-after-vaccine-administration.

## Results

Twelve healthy donors (67% female, mean age ± standard deviation 28.5±6.06 years) without pre-existing conditions and not undergoing any medication were included in the study. For a detailed description, see supplemental table 1. No adverse events were reported.

### Receptors and adhesion proteins expression in non-stimulated platelets

Mass cytometry acquired multidimensional relative protein expression quantification of 18 proteins (supplemental table 2) at single-cell level for each sample. To evaluate the platelet activation, we quantified the expression of five activation markers (P-Selectin, LAMP-1, LAMP-3, CD40L and PAC-1) before and after vaccination. The median signal intensity of all activation markers remained stable after vaccination and did not differ compared to pre-vaccination levels (P-Selectin *P*=0.169, LAMP-3 *P*=0.614 and PAC-1 *P*=0.746). Of note, the expression level at the membrane of CD40L and LAMP-1, normally stored within the platelets, were neither detectable before vaccination nor afterwards. Expression levels over time of P-Selectin and LAMP-3 are shown in Figure 1A. Median signal intensity of all activation markers are provided in supplemental table 2. We measured the expression levels of the following transmembrane receptors and adhesion proteins: CD41, CD61, CD42a, CD42b, PAR1, CD29, CD69, CD31, CD36, CD47, CD9, and CD40. Mass cytometry did not detect any differences before and after vaccination (Figure 1B, P values and median signal intensity provided in the supplemental table 2 and 3).

### Platelet expression after TRAP stimulation

To investigate platelet reactivity, we measured the activation marker expression after stimulation with TRAP. Protein expression rose after TRAP-stimulation similarly before and after vaccine administration (Figure 1C). The expression of constitutive transmembrane receptors and adhesion proteins after TRAP-stimulation did not differ before and after vaccination (supplemental table 3).

### Clustering analysis

The FlowSOM algorithm detected 8 clusters based on relative change in area under the cumulative distribution function curve. Activation marker expression (P-Selectin, LAMP-1, LAMP-3, CD40L and PAC-1) in the singular clusters did not change after vaccination (supplemental table 4). Figure 2A shows P-Selectin and LAMP-3 expression on FlowSOM for TP0, TP1 and TP4 as well as for the TRAP-stimulated TP0 samples. We provide further FlowSOM analysis in supplemental figure 2. Overall, no significant differences were detected within clusters concerning activation marker expression.

**Figure 2:**
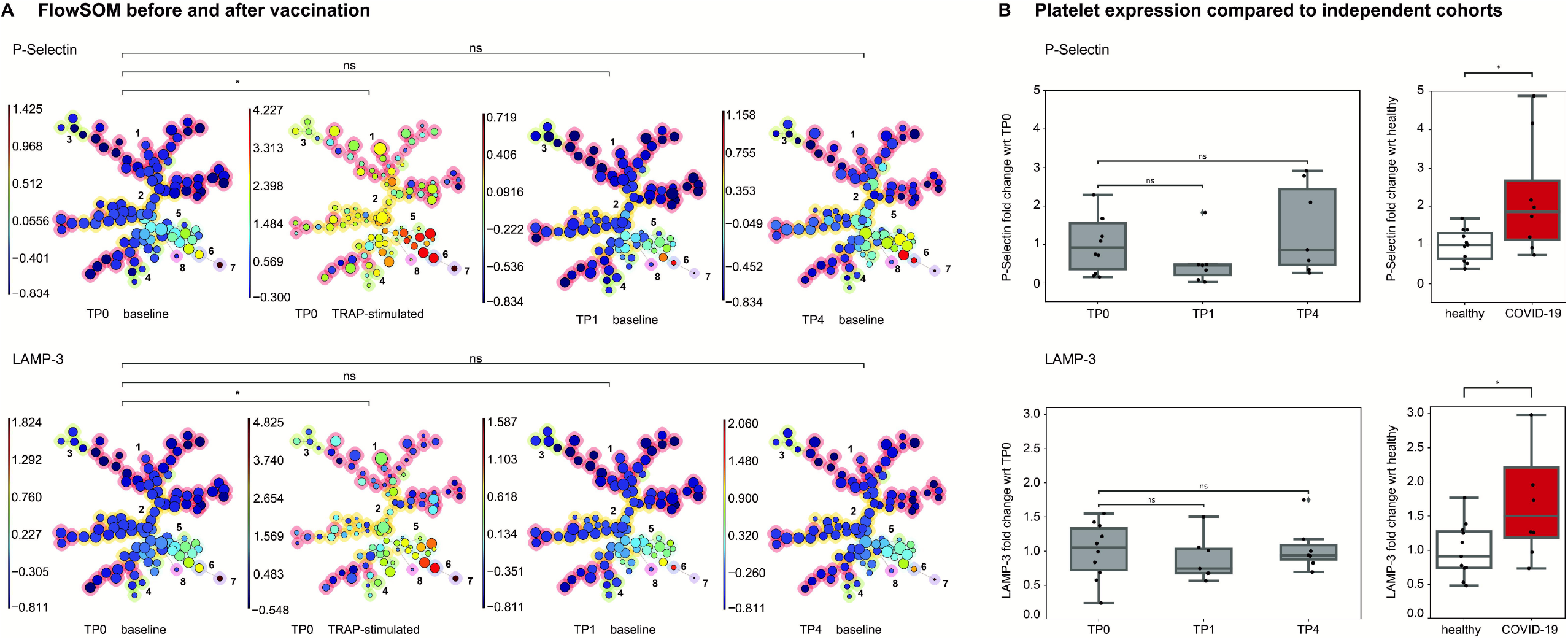
P-Selectin and LAMP-3 expression and comparison with COVID-19 patients. A) FlowSOM analysis of platelet samples. Platelets were clustered into SOM (self-organizing map) of clusters, which were then merged into 8 meta-clusters. The meta-clustering of the FlowSOM nodes is indicated by the background color of the nodes (red, orange, lime, green, cyan, violet, purple and magenta) and the numbers from 1-8. Separate FlowSOM trees show the P-Selectin (first row) and LAMP-3 (second row) expression across the clusters for non-stimulated platelets for TP0, TP1 and TP4 and after stimulation with 10 µM TRAP for TP0. All TPs were compared to TP0 (non-stimulated sample). A star (*) indicates a significant difference found in all clusters. ns= no significant in any of the clusters. Respective p values are shown in supplemental table 4. B) Platelet activation marker expression compared to an independent cohort of healthy donors and of COVID-19 patients. Fold change difference to control groups (TP0 and healthy controls) is shown. wrt= with reference to. TP0 n=10, TP1 n=8, TP4 n=7, ns=not significant.

### Platelet expression in comparison with independent cohorts

We compared protein expression levels before and after vaccination with two independent cohorts analyzed previously with the same platform^8^: COVID-19 symptomatic patients with no pre-existing condition requiring hospitalization and a healthy control group of donors tested negative for SARS-CoV-2 infection. Figure 2B provides an overview of P-Selectin and LAMP-3 expression. Expression levels of activation markers are comparable with those of healthy donors tested negative for COVID-19 and differ from COVID-19 patients.

## Discussion

Using mass cytometry, we investigated the expression of activation markers and regulatory transmembrane proteins in platelets of healthy donors undergoing BNT162b2 vaccination. The main finding of this study is that the exposition to spike protein upregulation through BNT162b2 does not alter the expression of the most important platelet adhesion proteins and receptors.

CyTOF allows the expression quantification of a large number of markers at single-cell levels at the same time. Avoiding the spectral limitation of flow-cytometry, this method allows a precise quantification of transmembrane proteins expression and it can be used to investigate the platelet expression phenotype with high-definition^20^. We here provide the first mass cytometric analysis of protein expression after SARS-CoV-2 vaccination.

A recent study suggested a direct effect on thrombocytes of the spike protein S, sought to be mediated through the ACE2 receptor^9^. Zang et al. reported that the spike protein alone (without presence of the virus) can dose-dependently enhance platelet aggregation, ATP release and dense granule secretion. Moreover, they reported that the spike protein induces GPIIb/IIIa activation and P-Selectin expression even in the absence of agonists^9^. In contrast to these findings, we observed that the expression level of the activated form of GPIIb/IIIa (PAC-1) as well as of other activation markers - including P-Selectin - does not change after the spike protein upregulation mediated by vaccination with BNT162b2 (Figure 1A-B and supplemental figure S1). Of note, the expression profiles after vaccination differed from COVID-19 patients characterized by the same experimental pipeline and remained similar to an independent control cohort (Figure 2B).

Platelet reactivity to physiological stimulus with TRAP was not altered after vaccination in contrast to the dysregulation observed during SARS-CoV-2 infection^8,21^. Clustering analysis of non-stimulated platelets did not detect a phenotypic modification after vaccination, in contrast to the specific cluster activation observed previously during SARS-CoV-2 infection^8^. Hence, our study shows that the vaccination does not generally lead to platelet activation or dysfunctional reactivity.

We acknowledge the limitations of our investigations: with our restricted sample size we did non aim to investigate the phenotypic expression of rare complications such as thromboembolic events or the presence of vaccine-induced immune thrombotic thrombocytopenia (VITT), that has been reported after ChAdOx1 administration^15,22^. Moreover, we recruited donors undergoing vaccination only with BNT162b2 administration. While all vaccines induce protein spike S upregulation, they use different vectors. Thus, our findings cannot be directly translated to all available vaccines. Nevertheless, our study provides the first phenotypic characterization of platelets of healthy donors undergoing vaccination and provides additional biomolecular information. While a direct activity of spike protein S on platelets cannot be excluded, our data suggest that the spike protein S expression levels mediated by SARS-CoV-2 vaccines do not significantly alter platelet biology and function.

In conclusion, mass cytometry of platelets after BNT162b2 vaccination did not detect any alteration of the platelet expression profile and platelet reactivity. Further studies are needed to characterize the pathophysiology of the rare thromboembolic events occurring after SARS-CoV-2 vaccination.

## Data Availability

All mass cytometry data and FlowSOM results have been made available at flowrepository.org and can be accessed at repository ID FR-FCM-Z3XS. The scripts used in this analysis have been deposited at github.com and can be accessed at https://github.com/biomedbigdata/Platelet-activation-after-vaccine-administration.

https://flowrepository.org/id/FR-FCM-Z3XS

https://github.com/biomedbigdata/Platelet-activation-after-vaccine-administration

## Acknowledgment

The authors would like to express their gratitude to the health workers of the University hospital rechts der Isar for their support regardless of extremely challenging working conditions due to the SARS-CoV-2 outbreak. The authors thank the Core Facility Cell Analysis at TranslaTUM (CFCA), Klinikum rechts der Isar of Technical University Munich for their support during this study. The graphical abstract was created with biorender.com.

## What is known on this topic?

- SARS-CoV-2 spike protein enhances thrombosis formation, stimulates human platelets to release pro-coagulant factors and promotes platelet-leukocyte aggregate formation even in absence of the virus.
- Platelet expression and reactivity after SARS-CoV-2 vaccination have not been investigated yet.

## What does this paper add?

- BNT162b2 vaccination does not alter the expression of platelet adhesion proteins and receptors
- Vaccine-induced spike protein S upregulation does not lead to platelet activation or dysfunction.
- Cluster analysis excluded a dysregulation of platelet subgroups.

## References

1. Wichmann D, Sperhake J-P, Lütgehetmann M et al. Autopsy Findings and Venous Thromboembolism in Patients With COVID-19: A Prospective Cohort Study. Ann Intern Med. 2020;173:268–277.

2. Poissy J, Goutay J, Caplan M et al. Pulmonary Embolism in COVID-19 Patients: Awareness of an Increased Prevalence. Circulation. 2020;142:184–186.

3. Bangalore S, Sharma A, Slotwiner A et al. ST-Segment Elevation in Patients with Covid-19 — A Case Series. New Engl J Med. 2020;382:2478–2480.

4. Leentjens J, Haaps TF van, Wessels PF, Schutgens REG, Middeldorp S. COVID-19-associated coagulopathy and antithrombotic agents—lessons after 1 year. Lancet Haematol. 2021;

5. Manne BK, Denorme F, Middleton EA et al. Platelet Gene Expression and Function in COVID-19 Patients. Blood. 2020;

6. Nicolai L, Leunig A, Brambs S et al. Immunothrombotic Dysregulation in COVID-19 Pneumonia is Associated with Respiratory Failure and Coagulopathy. Circulation. 2020;

7. Zaid Y, Puhm F, Allaeys I et al. Platelets Can Associate With SARS-CoV-2 RNA and Are Hyperactivated in COVID-19. Circ Res. 2020;127:1404–1418.

8. Bongiovanni D, Klug M, Lazareva O et al I. SARS-CoV-2 infection is associated with a pro-thrombotic platelet phenotype. Cell Death Dis. 2021;12:50.

9. Zhang S, Liu Y, Wang X et al. SARS-CoV-2 binds platelet ACE2 to enhance thrombosis in COVID-19. J Hematol Oncol. 2020;13:120.

10. Polack FP, Thomas SJ, Kitchin N et al. Safety and Efficacy of the BNT162b2 mRNA Covid-19 Vaccine. New Engl J Med. 2020;383:2603–2615.

11. Baden LR, Sahly HME, Essink B et al. Efficacy and Safety of the mRNA-1273 SARS-CoV-2 Vaccine. New Engl J Med. 2020;

12. Voysey M, Clemens SAC, Madhi SA et al. Safety and efficacy of the ChAdOx1 nCoV-19 vaccine (AZD1222) against SARS-CoV-2: an interim analysis of four randomised controlled trials in Brazil, South Africa, and the UK. Lancet. 2021;397:99–111.

13. Castelli GP, Pognani C, Sozzi C, Franchini M, Vivona L. Cerebral venous sinus thrombosis associated with thrombocytopenia post-vaccination for COVID-19. Crit Care. 2021;25:137.

14. Carli G, Nichele I, Ruggeri M, Barra S, Tosetto A. Deep vein thrombosis (DVT) occurring shortly after the second dose of mRNA SARS-CoV-2 vaccine. Intern Emerg Med. 2021;16:803–804.

15. Greinacher A, Thiele T, Warkentin TE, Weisser K, Kyrle PA, Eichinger S. Thrombotic Thrombocytopenia after ChAdOx1 nCov-19 Vaccination. New Engl J Med. 2021;

16. Bernlochner I, Klug M, Larasati D et al. Sorting and magnetic-based isolation of reticulated platelets from peripheral blood. Platelets. 2020;1–7.

17. Kotecha N, Krutzik PO, Irish JM. Web-Based Analysis and Publication of Flow Cytometry Experiments. Curr Protoc Cytom. 2010;53:10.17.1-10.17.24.

18. Harris CR, Millman KJ, Walt SJ van der et al. Array programming with NumPy. Nature. 2020;585:357–362.

19. Spidlen J, Breuer K, Rosenberg C, Kotecha N, Brinkman RR. FlowRepository: A resource of annotated flow cytometry datasets associated with peer-reviewed publications. Cytom Part A. 2012;81A:727–731.

20. Blair TA, III ALF. Platelet surface marker analysis by mass cytometry. Platelets. 2019;31:1–8.

21. Clark CC, Jukema BN, Barendrecht AD et al. Thrombotic Events in COVID-19 Are Associated With a Lower Use of Prophylactic Anticoagulation Before Hospitalization and Followed by Decreases in Platelet Reactivity. Frontiers Medicine. 2021;8:650129.

22. Hundelshausen P von, Lorenz R, Siess W, Weber C. Vaccine-induced immune thrombotic thrombocytopenia (VITT): targeting pathomechanisms with Bruton tyrosine kinase inhibitors. Thromb Haemostasis. 2021;

