## Supplemental Material for "Platelet expression and reactivity after BNT162b2 vaccine administration"

Running title: SARS-CoV-2 vaccination and platelet expression

Melissa Klug M.Sc*[,1,2,3], Olga Lazareva M.Sc*[,3], Kilian Kirmes B.Sc[1], Marc Rosenbaum PhD[4,5], Marina Lukas M.D[6], Simon Weidlich M.D.[7], Christoph D. Spinner M.D.[7], Moritz von Scheidt M.D.[2,8], Rosanna Gosetti M.D.[1], Jan Baumbach PhD[9], Jürgen Ruland M.D[4,5], Gianluigi Condorelli M.D.[10], Karl-Ludwig Laugwitz M.D.[1,2], Markus List Ph.D. [†,3] Isabell Bernlochner M.D.[†,1,2] and Dario Bongiovanni M.D.[†,1,2,10]
* equal contribution
† these authors are joint senior author

[1] Department of Internal Medicine I, School of Medicine, University hospital rechts der Isar, Technical University of Munich, Munich, Germany;
[2] German Center for Cardiovascular Research (DZHK), Partner Site Munich Heart Alliance, Munich, Germany;
[3] Chair of Experimental Bioinformatics, TUM School of Life Sciences Weihenstephan, Technical University of Munich, Munich, Germany;
[4] School of Medicine, Institute of Clinical Chemistry and Pathobiochemistry, Technical University of Munich, Munich, Germany;
[5] TranslaTUM, Center for Translational Cancer Research, Technical University of Munich, Munich, Germany;
[6] Department of Internal Medicine III, School of Medicine, University hospital rechts der Isar, Technical University of Munich, Munich, Germany;
[7] Department of Internal Medicine II, School of Medicine, University hospital rechts der Isar, Technical University of Munich, Munich, Germany;
[8] Deutsches Herzzentrum München, Cardiology, Technical University of Munich, Munich, Germany;
[9] Chair of Computational Systems Biology, University of Hamburg, Hamburg, Germany
[10] Department of Cardiovascular Medicine, Humanitas Clinical and Research Center IRCCS and Humanitas University, Rozzano, Milan, Italy

### <u>Supplemental Material</u>

**Supplementary Tables** **page 2**

**Supplementary Figures** **page 11**

**Major Resources Table** **page 13**

**Supplementary Table 1.** Baseline Characteristics of healthy donors.

| **Donor** | **Gender** | **Age** |
|---|---|---|
| 1 | female | 31-36 |
| 2 | female | 25-30 |
| 3 | female | 31-36 |
| 4 | female | 25-30 |
| 5 | female | 43-48 |
| 6 | male | 25-30 |
| 7 | male | 31-36 |
| 8 | male | 19-24 |
| 9 | female | 25-30 |
| 10 | female | 25-30 |
| 11 | female | 25-30 |
| 12 | male | 19-24 |

**Supplementary Table 2.** Median signal intensity of baseline (non-stimulated) and 10 µM TRAP stimulated (activated) samples at timepoints TP0 until TP5.

| **Target antigen** | **TRAP activation** | **TP0** | **TP1** | **TP2** | **TP3** | **TP4** | **TP5** |
|---|---|---|---|---|---|---|---|
| P-Selectin | baseline | 0.4 | 0.2 | 0.37 | 0.47 | 0.37 | 0.33 |
| P-Selectin | activated | 3.92 | 4.12 | 3.88 | 4.4 | 4.11 | 3.62 |
| PAR1 | baseline | 1.35 | 1.35 | 1.46 | 1.35 | 1.44 | 1.38 |
| PAR1 | activated | 1.32 | 1.4 | 1.37 | 1.39 | 1.43 | 1.35 |
| PAC1 | baseline | 0.52 | 0.67 | 0 | 0.54 | 0.31 | 0.12 |
| PAC1 | activated | 1.74 | 1.89 | 1.25 | 1.33 | 1.19 | 1.37 |
| LAMP3 | baseline | 0.27 | 0.19 | 0.21 | 0.29 | 0.24 | 0.21 |
| LAMP3 | activated | 1.65 | 1.37 | 1.62 | 1.88 | 1.58 | 1.01 |
| LAMP1 | baseline | 0 | 0 | 0 | 0 | 0 | 0 |
| LAMP1 | activated | 0.05 | 0.11 | 0 | 0.43 | 0.25 | 0 |
| CD9 | baseline | 4.16 | 4.04 | 4.18 | 4.46 | 4.27 | 4.03 |
| CD9 | activated | 4.26 | 4.29 | 4.34 | 4.52 | 4.29 | 4.23 |
| CD69 | baseline | 1.65 | 1.62 | 1.77 | 1.77 | 1.72 | 1.65 |
| CD69 | activated | 1.98 | 2.11 | 2.1 | 2.3 | 2.18 | 2.1 |
| CD61 | baseline | 2.13 | 2.08 | 2.01 | 2.34 | 2.09 | 1.84 |
| CD61 | activated | 2.27 | 2.27 | 2.33 | 2.39 | 2.25 | 2.08 |

| | | | | | | | |
|---|---|---|---|---|---|---|---|
| CD47 | baseline | 2.04 | 1.92 | 1.94 | 2.26 | 2.12 | 1.82 |
| CD47 | activated | 2.14 | 2.18 | 2.25 | 2.36 | 2.22 | 2.05 |
| CD42b | baseline | 3 | 2.93 | 2.82 | 3.17 | 3.08 | 2.89 |
| CD42b | activated | 2.92 | 2.98 | 2.98 | 3.16 | 3.18 | 3.06 |
| CD42a | baseline | 3.49 | 3.34 | 3.53 | 3.74 | 3.52 | 3.3 |
| CD42a | activated | 3.45 | 3.44 | 3.6 | 3.66 | 3.54 | 3.33 |
| CD41 | baseline | 1.35 | 1.29 | 1.24 | 1.61 | 1.38 | 1.06 |
| CD41 | activated | 1.45 | 1.45 | 1.44 | 1.65 | 1.44 | 1.22 |
| CD40L | baseline | 0 | 0 | 0 | 0 | 0 | 0 |
| CD40L | activated | 0.05 | 0 | 0.01 | 0.06 | 0.1 | 0 |
| CD40 | baseline | 0 | 0 | 0 | 0.02 | 0 | 0 |
| CD40 | activated | 0 | 0.03 | 0.04 | 0.09 | 0.05 | 0.03 |
| CD36 | baseline | 2.79 | 2.64 | 2.85 | 2.91 | 2.72 | 2.59 |
| CD36 | activated | 2.94 | 2.83 | 2.87 | 3.04 | 3 | 2.81 |
| CD31 | baseline | 1.37 | 1.17 | 1.28 | 1.52 | 1.33 | 1.07 |
| CD31 | activated | 1.45 | 1.47 | 1.46 | 1.65 | 1.55 | 1.34 |
| CD3 | baseline | 0 | 0 | 0 | 0 | 0 | 0 |
| CD3 | activated | 0 | 0 | 0 | 0 | 0 | 0 |
| CD29 | baseline | 3.71 | 3.59 | 3.67 | 4.03 | 3.86 | 3.52 |

| | | | | | | | |
|---|---|---|---|---|---|---|---|
| CD29 | activated | 3.77 | 3.75 | 3.84 | 4.06 | 3.9 | 3.72 |

**Supplementary Table 3.** Kruskal-Wallis p-values for multi-group comparison between timepoints TP0 until TP5 for baseline (non-stimulated) and 10 μM TRAP stimulated (activated) samples.

| **Marker** | **TRAP activation** | **p-value** |
|---|---|---:|
| P-Selectin | baseline | 0.169 |
| P-Selectin | activated | 0.937 |
| PAR1 | baseline | 0.118 |
| PAR1 | activated | 0.342 |
| PAC1 | baseline | 0.746 |
| PAC1 | activated | 0.547 |
| LAMP3 | baseline | 0.614 |
| LAMP3 | activated | 0.577 |
| LAMP1[1] | baseline | 1 |
| LAMP1 | activated | 0.085 |
| CD9 | baseline | 0.726 |
| CD9 | activated | 0.911 |
| CD69 | baseline | 0.194 |
| CD69 | activated | 0.501 |
| CD61 | baseline | 0.641 |

[1] Markers with 0 median expression that did not change over time have p-value equal to 1

| | | |
|---|---|---|
| CD61 | activated | 0.972 |
| CD47 | baseline | 0.88 |
| CD47 | activated | 0.82 |
| CD42b | baseline | 0.442 |
| CD42b | activated | 0.342 |
| CD42a | baseline | 0.093 |
| CD42a | activated | 0.651 |
| CD41 | baseline | 0.675 |
| CD41 | activated | 0.972 |
| CD40L | baseline | 1 |
| CD40L | activated | 0.641 |
| CD40 | baseline | 0.671 |
| CD40 | activated | 0.294 |
| CD36 | baseline | 0.332 |
| CD36 | activated | 0.653 |
| CD31 | baseline | 0.53 |
| CD31 | activated | 0.609 |
| CD3 | baseline | 1 |
| CD3 | activated | 1 |

| | | |
|---|---|---|
| CD29 | baseline | 0.82 |
| CD29 | activated | 0.864 |

**Supplementary Table 4**: Comparison of activation marker expression in different FlowSOM clusters using Mann–Whitney U test (one-sided p-values).

| **Marker** | **Cluster** | **TP0 to TP1 baseline** | **TP0 to TP4 baseline** | **TP0 baseline to TP0 stimulated** |
|---|---|---|---|---|
| P-Selectin | 1 | 0.90779236 | 0.21606608 | 0.0000608 |
| P-Selectin | 2 | 0.74741987 | 0.11125611 | 0.0000621 |
| P-Selectin | 3 | 0.84025828 | 0.42093366 | 0.0000608 |
| P-Selectin | 4 | 0.86664138 | 0.20340651 | 0.0000621 |
| P-Selectin | 5 | 0.84656264 | 0.15275354 | 0.0000621 |
| P-Selectin | 6 | 0.48228235 | 0.07852617 | 0.0000621 |
| P-Selectin | 7 | 0.44698657 | 0.36633913 | 0.03627463 |
| P-Selectin | 8 | 0.34463884 | 0.24712999 | 0.0001882 |
| PAC1 | 1 | 0.32545115 | 0.6910271 | 0.0182663 |
| PAC1 | 2 | 0.25147419 | 0.48052941 | 0.01575626 |
| PAC1 | 3 | 0.24619094 | 0.59979009 | 0.01780541 |
| PAC1 | 4 | 0.3116322 | 0.63397704 | 0.01324732 |
| PAC1 | 5 | 0.34463884 | 0.51945867 | 0.01888567 |
| PAC1 | 6 | 0.34463884 | 0.07852617 | 0.00756157 |
| PAC1 | 7 | 0.09881055 | 0.6697254 | 0.0650149 |
| PAC1 | 8 | 0.36107275 | 0.75287001 | 0.00241898 |

| | | | | |
|---|---|---|---|---|
| LAMP3 | 1 | 0.65536116 | 0.55819127 | 0.0000621 |
| LAMP3 | 2 | 0.28178731 | 0.26293109 | 0.0000621 |
| LAMP3 | 3 | 0.87609331 | 0.76844201 | 0.0000621 |
| LAMP3 | 4 | 0.84656264 | 0.40362508 | 0.0000621 |
| LAMP3 | 5 | 0.62209329 | 0.20340651 | 0.0000621 |
| LAMP3 | 6 | 0.28178731 | 0.20340651 | 0.0000825 |
| LAMP3 | 7 | 0.44698657 | 0.51945867 | 0.02237965 |
| LAMP3 | 8 | 0.22505105 | 0.05367327 | 0.00052937 |
| LAMP1 | 6 | 0.40949594 | 0.14745665 | 0.0000785 |
| LAMP1 | 7 | 0.11516553 | 0.3302746 | 0.04898049 |
| LAMP1 | 8 | 0.84284767 | 0.83051008 | 0.00441037 |
| CD40L | 6 | 0.84713361 | 0.10621535 | 0.000059 |
| CD40L | 7 | 0.41192703 | 0.5583334 | 0.0565511 |
| CD40L | 8 | 0.43537817 | 0.83051008 | 0.0117525 |

### Supplementary Figures

### Supplementary Figure S I

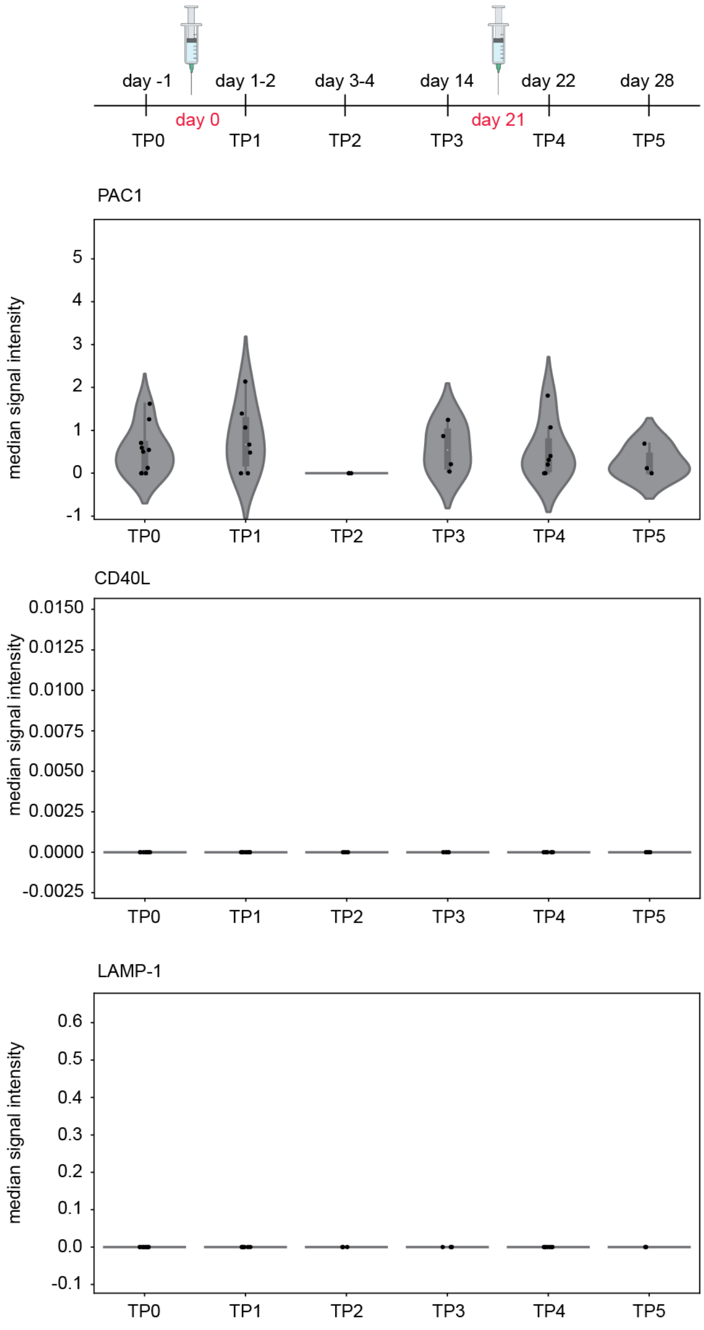


**Supplementary Figure S I:** Activation marker expression at baseline level. PAC1, CD40L and LAMP-1 median signal intensity is shown at the different timepoints. Each dot represents the median signal intensity of one sample. Changes in expression were found insignificant between timepoints (Kruskal-Wallis test, Supplementary table 3). TP0 n=10, TP1 n=8, TP 2=3, TP 3 n=4, TP4 n=7, TP5 n=3.

**Supplementary Figure S II**

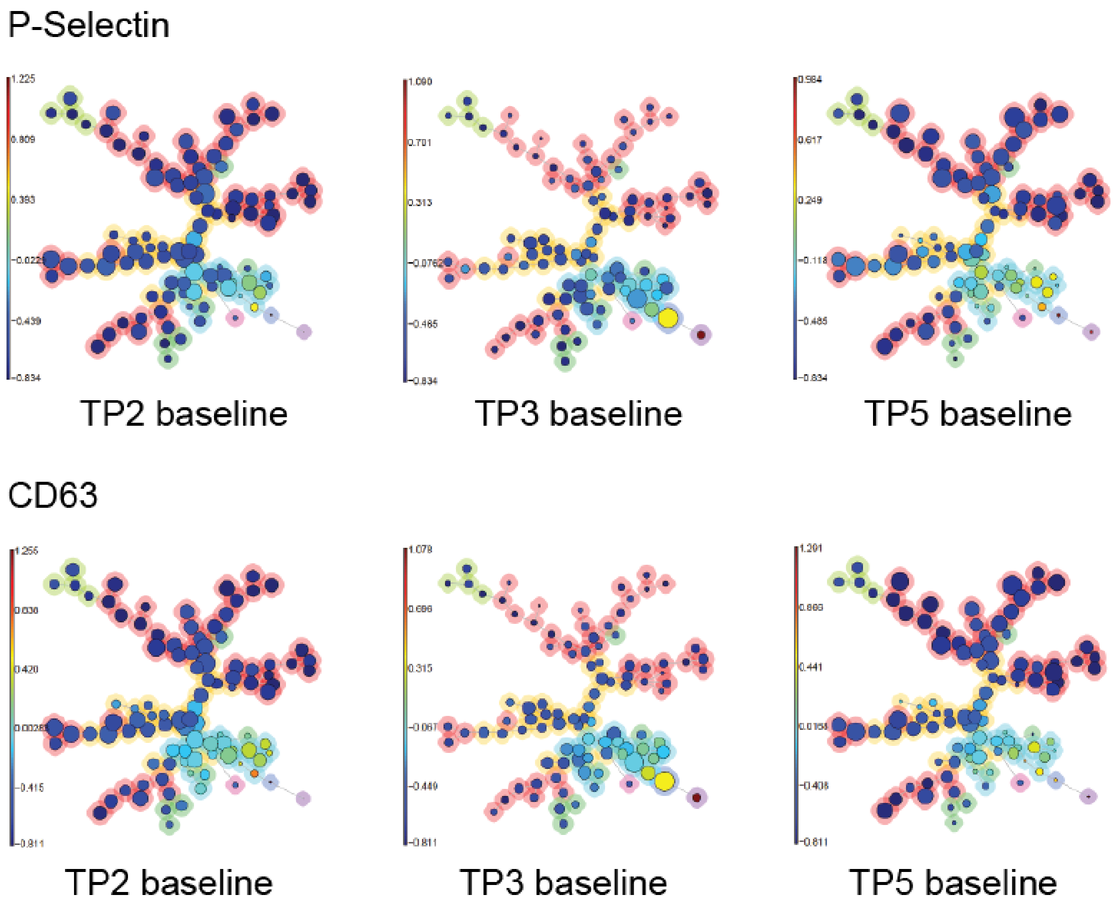


**Supplementary Figure S II FlowSOM analysis of TP2, TP3 and TP5 samples:** Platelets were clustered into SOM (self-organizing map) of clusters, which were then merged into 8 meta-clusters. The meta-clustering of the FlowSOM nodes is indicated by the background color of the nodes (red, orange, lime, green, cyan, violet, purple and magenta) and the numbers from 1-8. Separate FlowSOM trees show the P-Selectin (first row) and LAMP-3 (second row) expression across the clusters for non- stimulated platelets for TP2, TP3 and TP5., TP 2 n=3, TP 3 n=4 and TP5 n=3.

<u>**Major Resources Table**</u>

**Antibodies**

| **Target antigen** | **Vendor or Source** | **Catalog #** | **Working concentration** | **Persistent ID / URL** |
|---|---|---|---|---|
| CD62P | Thermofisher | MA1-81809 | 0.5 mg/ml | https://www.thermofisher.com/antibody/product/P-Selectin-Antibody-clone-Psel-KO-2-7-Monoclonal/MA1-81809 |
| PAR1 | Thermofisher | 35-2200 | 0.5 mg/ml | https://www.thermofisher.com/antibody/product/PAR1-Antibody-clone-ATAP2-Monoclonal/35-2200 |
| CD42a | Thermofisher | MA1-91023 | 0.5 mg/ml | https://www.thermofisher.com/antibody/product/CD42a-Antibody-clone-GR-P-Monoclonal/MA1-91023 |
| Activated GPIIbIIIa / αIIbβ3 (PAC-1) | BD Biosciences | 340535 | 0.5 mg/ml | https://www.bdbiosciences.com/us/reagents/research/clinical-research---ruo-gmp/purified-antibodies/purified-mouse-anti-human-pac-1-pac-1/p/340535 |
| CD107a/ LAMP1 | Fluidigm Sciences | 3151002B | 0.5 mg/ml | https://www.fluidigm.com/reagents/proteomics/3151002b-antihuman-cd107a-lamp1--h4a3--151eu--100tests |
| CD154/ CD40L | Fluidigm Sciences | 3168006B | 0.5 mg/ml | https://www.fluidigm.com/reagents/proteomics/3168006b-antihuman-cd154-cd40l--24-31--168er--100tests |

| | | | | |
|---|---|---|---|---|
| CD29 | Fluidigm Sciences | 3156007B | 0.5 mg/ml | https://www.fluidigm.com/reagents/proteomics/3156007b-antihuman-cd29--ts2-16--156gd--100tests |
| CD3 | Fluidigm Sciences | 3170001B | 0.5 mg/ml | https://www.fluidigm.com/reagents/proteomics/3170001b-antihuman-cd3--ucht1--170er--100tests |
| CD31/ PECAM | Fluidigm Sciences | 3145004B | 0.5 mg/ml | https://www.fluidigm.com/reagents/proteomics/3145004b-antihuman-cd31-pecam-1--wm59--145nd--100tests |
| CD36 | Fluidigm Sciences | 3152007B | 0.5 mg/ml | https://www.fluidigm.com/reagents/proteomics/3152007b-antihuman-cd36--5-271--152sm--100tests |
| CD40 | Fluidigm Sciences | 3142010B | 0.5 mg/ml | https://www.fluidigm.com/reagents/proteomics/3142010b-antihuman-cd40--5c3--142nd--100tests |
| CD41 | Fluidigm Sciences | 3089004B | 0.5 mg/ml | https://www.fluidigm.com/reagents/proteomics/3089004b-antihuman-cd41--hip8--89y--100tests |
| CD42b | Fluidigm Sciences | 3144020B | 0.5 mg/ml | https://www.fluidigm.com/reagents/proteomics/3144020b-antihuman-cd42b--hip1--144nd--100tests |
| CD47 | Fluidigm Sciences | 3209004B | 0.5 mg/ml | https://www.fluidigm.com/reagents/proteomics/3209004b-antihuman-cd47--cc2c6--209bi--100tests |

| | | | | |
|---|---|---|---|---|
| CD61 | Fluidigm Sciences | 3146011B | 0.5 mg/ml | https://www.fluidigm.com/reagents/proteomics/3146011b-antihuman-cd61--vi-pl2--146nd--100tests |
| CD63 | Fluidigm Sciences | 3150021B | 0.5 mg/ml | https://www.fluidigm.com/reagents/proteomics/3150021b-antihuman-cd63--h5c6--150nd--100tests |
| CD69 | Fluidigm Sciences | 3162001B | 0.5 mg/ml | https://www.fluidigm.com/reagents/proteomics/3162001b-antihuman-cd69--fn50--162dy--100tests |
| CD9 | Fluidigm Sciences | 3171009B | 0.5 mg/ml | https://www.fluidigm.com/reagents/proteomics/3171009b-antihuman-cd9--sn4-c3-3a2--171yb--100tests |

**Data & Code Availability**

| **Description** | **Source / Repository** | **Persistent ID / URL** |
|---|---|---|
| Mass Cytometry data, accession under FR-FCM-Z3XS. | flowrepository.org | https://flowrepository.org/id/FR-FCM-Z3XS |
| Scripts used in the analysis | Python and R script/github | https://github.com/biomedbigdata/Platelet-activation-after-vaccine-administration |

| **Reagents** | **Source / Repository** | **Persistent ID / URL** |
|---|---|---|

| | | |
|---|---|---|
| Maxpar® X8 Multimetal Labeling Kit | Fluidigm Sciences | 201300/ https://www.fluidigm.com/reagents/proteomics/201300-maxpar-x8-multimetal-labeling-kit--40rxn |
| Cell-ID™ Intercalator-Ir | Fluidigm Sciences | 201192A/ https://www.fluidigm.com/reagents/proteomics/201192a-cellid-intercalator-ir--125micrometer |
| Cell-ID™ Cisplatin | Fluidigm Sciences | 201064/ https://www.fluidigm.com/reagents/proteomics/201064-cellid-cisplatin--100microliter |
| Maxpar® Cell Staining Buffer | Fluidigm Sciences | 201068/ https://www.fluidigm.com/reagents/proteomics/201068-maxpar-cell-staining-buffer--500ml |
| Maxpar® Fix and PermBuffer | Fluidigm Sciences | 201067/ https://www.fluidigm.com/reagents/proteomics/201067-maxpar-fix-and-perm-buffer--100ml |
| Maxpar® Cell Acquisition Solution | Fluidigm Sciences | https://www.fluidigm.com/reagents/proteomics/201240-maxpar-cell-acquisition-solution%E2%80%94200-ml |
| AB Stabilizer, PBS base | Boca Scientific | 131050/ https://www.bocascientific.com/antibody-stabilizer-p-4369.html |
